# Role of IgG against N-protein of SARS-CoV2 in COVID19 clinical outcomes

**DOI:** 10.1101/2020.09.23.20197251

**Authors:** Mayank Batra, Runxia Tian, Chongxu Zhang, Emile Clarence, Camila Sofia Sacher, Justin Nestor Miranda, Justin Rafa O De La Fuente, Megan Mathew, Desmond Green, Sayari Patel, Maria Virginia Perez Bastidas, Sara Haddadi, Mukunthan Murthi, Miguel Santiago Gonzalez, Shweta Kambali, Kayo H M Santos, Huda Asif, Farzaneh Modarresi, Mohammad Faghihi, Mehdi Mirsaeidi

**Affiliations:** Division of Pulmonary and Critical Care, University of Miami, Miami, FL; School of Medicine, University of Miami, Miami, FL; Express Gene Laboratory, Miami, FL

**Keywords:** IgG, SARS-CoV2, Nucleoprotein, COVID19

## Abstract

The Nucleocapsid Protein (N Protein) of severe acute respiratory syndrome Coronavirus 2 (SARS-CoV2) is located in the viral core. Immunoglobulin G (IgG) targeting N protein is detectable in the serum of infected patients. The effect of high titers of IgG against N-protein on clinical outcomes of SARS-CoV2 disease has not been described. We studied 400 RT-PCR confirmed SARS-CoV2 patients to determine independent factors associated with poor outcomes, including MICU admission, prolonged MICU stay and hospital admissions, and in-hospital mortality. We also measured serum IgG against the N protein and correlated its concentrations with clinical outcomes. We found that several factors, including Charlson comorbidity Index (CCI), high levels of IL6, and presentation with dyspnea were associated with poor clinical outcomes. It was shown that higher CCI and higher IL6 levels were independently associated with in-hospital mortality. Anti-N protein IgG was detected in the serum of 55 (55%) patients at the time of admission. A high concentration of antibodies, defined as signal to cut off ratio (S/Co)> 1.5 (75 percentile of all measurements), was found in 25 (25%) patients. The multivariable logistic regression models showed that between being an African American, higher CCI, lymphocyte counts, and S/Co ratio> 1.5, only S/Co ratio were independently associated with MICU admission and longer length of stay in hospital. This study recommends that titers of IgG targeting N-protein of SARS-CoV2 at admission is a prognostic factor for the clinical course of disease and should be measured in all patients with SARS-CoV2 infection.

## Introduction

Severe acute respiratory syndrome coronavirus 2 (SARS-CoV-2) and its resultant disease, Coronavirus Disease 2019 (COVID19), has spread rapidly since its first report in Wuhan, China, to become a public health threat of global importance ^1,2^. According to the World Health Organization, there are over 30 million cases worldwide as of September 16^th^ 2020, with the United States accounting for 6.6 million cases and over 196,000 deaths alone. This prolific virus is a positive-sense single-stranded RNA belonging to the Betacoronavirus genus.

SARS-CoV-2 has four key structural proteins. The Nucleocapsid Protein (N Protein) is found in the viral core^3^. The Spike Protein (S Protein), Matrix Protein (M Protein) and Envelope Protein (E Protein)^3^ are all found on the virus’s outer surface. The S Protein is a key determinant in the viral host range and possibly infectivity^4^. Immunoglobulin G (IgG) developed against the S protein is believed to be a neutralizing antibody immune response and is currently the primary target for COVID19 vaccine trials. The M protein is the most abundant protein on the viral surface and is believed to play a role in viral budding from the host cell membrane^5,6^. The E protein is the smallest protein whose function is poorly defined, but it is thought to contribute to intracellular viral trafficking and viral protein assembly^7^.

Although the functionality of the N protein is not well understood, it has been well studied in SARS-CoV1 ^8^. The N protein is closely associated with viral RNA structure and function and plays a vital role in viral transcription and assembly while functioning as an RNA chaperone^9^. The N segment of the RNA has 3 distinct regions; a motif responsible for length of amino acids of N protein, a sumoylation motif (lysin residue), and a serin residue responsible for phosphorylation by a cyclin-dependent kinase complex ^8^.

A neutralizing antibody (Ab) may block interaction of a virus protein with the host receptor ^10^. However, a non-neutralizing Ab may even complicate the pathogenesis of some viral infections by antibody dependent enhancement (ADE). ADE is a phenomenon that is well described in viral infections, including Coronavirus ^11^. ADE represents a paradoxical Ab effect where the virus uses the antibodies to gain access to host immune cells. Specifically, the virus uses non-neutralizing Ab to attach to immune cells, replicate within macrophages and cytotoxic T cells, and consequently worsen the infection through cytotoxic effects on the host immune cells ^12^.

ADE is also associated with higher viremia ^13^. Non-neutralized viral particles may also lead to the extensive release of pro-inflammatory cytokines and the inhibition of anti-inflammatory cytokines causing immunopathology ^12^.

An ongoing debate in the medical community concerns the impact of ADE on SARS-CoV-2 infection and if ADE is contributing to the current pandemic. One possible factor leading to the trend of poorer prognosis with age may be previous exposure to various strains of Coronavirus, making the host more susceptible to ADE.

The N protein of SARS-CoV2 has high homology with other highly pathogenic members of the coronavirus family, with a molecular weight of about 46-kDa ^14^. Previous exposure could lead to the production of non-neutralizing Abs and establish an overwhelming pro-inflammatory state through ADE^15^. Patients who have higher levels of non-neutralizing IgG early in their disease may experience a worse infection due to ADE.

We hypothesize that IgG Abs against the N protein may be associated with the ADE phenomenon and higher levels of viremia. Elevated levels of coronavirus anti-N protein IgG may be associated with poorer outcomes such as increased length of hospitalization, increased likelihood of ICU admission, increased length of ICU admission, and increased mortality during hospitalization. To test these hypotheses, we conducted a prospective study on RT-PCR-confirmed COVID19 patients admitted to the University of Miami Hospital to determine clinical factors associated with poor outcomes. In addition, this study aimed to determine the association between serum SARS-CoV2 anti-N protein IgG levels and clinical outcomes of COVID19 disease. Further, we performed RT-PCR on RNA extracted from the whole blood of subjects with negative, low, and high concentrations of IgG for N-protein to detect viremia.

## Materials and Methods

### Ethics Statement

This study was approved by the Institutional Review Board (IRB) of the University of Miami.

### Study population

Patients who were seen in the emergency room (ER) or admitted to COVID19 units of University of Miami Hospital from March to the end of June 2020 were consented and enrolled in this study.

### Study design and sampling of peripheral blood

The study population included individuals admitted to the University of Miami Hospital with COVID19 infection confirmed via RT-PCR from nasopharyngeal swabs. Four hundred RT-PCR confirmed COVID19 patients were enrolled. Out of the 400 COVID-19 patients, 100 were consented to participate in the COVID19 biobank. Figure 1 shows the study algorithm. Serum samples were used to measure IgG against N-protein and whole blood was used for SARS-CoV2 RT-PCR.

**Figure 1.**
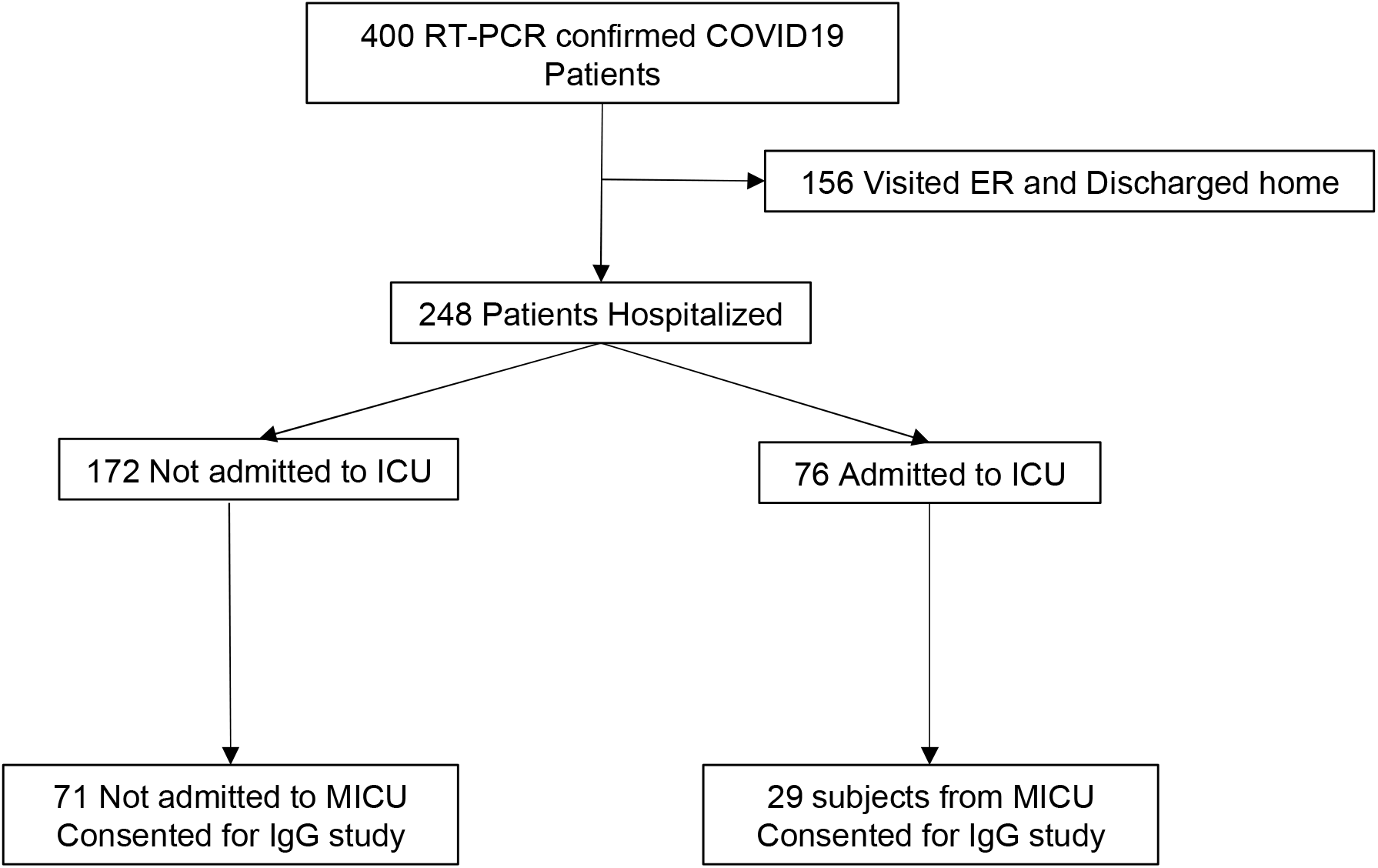
Shows the study algorithm.

All 100 serum samples were enrolled for IgG measurement. The single to cut off (S/Co) ratio was measured for all samples.

Fifteen patients were randomly selected for whole blood RNA extraction. Five patients with high IgG titers (defined as S/Co ratio >1.5), 5 with positive IgG but S/Co ratio<1.5, and 5 with undetectable IgG levels were selected at enrollment.

Peripheral blood samples were collected following enrollment and processed by centrifugation at 1800 g for 30 minutes. Serum and whole blood were aliquoted into 2 ml cryogenic tubes and stored at –80 C immediately for later analysis.

### Data collection and outcomes

Demographic, clinical, and outcome data were collected from the electronic medical record system for each patient. Charlson Comorbidity Index (CCI) was calculated using the comorbidities and age data for each patient. The main outcomes were ICU admission, length of stay in ICU, and in-hospital mortality. In-hospital mortality was defined as death occurring during the hospital stay.

### ELISA test

Serum samples were processed and analyzed for SARS-CoV-2 IgG antibodies and anti N-protein IgG levels using an ELISA kit for IgG by Epitope Diagnostics ^16^ according to manufacturer’s protocol. Cut off values for positive tests were determined by the manufacturer’s formula [1.1 X (xNC + 0.18)].^17^ Signal to cut-off ratio was calculated for each sample.

### RNA Isolation

RNA isolation from whole blood samples was performed by using a kit from ZymoResearch (Orange, CA, USA) according to the manufacturer’s recommendations.

### RT-PCR

The Express Gene SARS-CoV2 RT-PCR Diagnostic Panel was used to detect SARS C on extracted RNA from blood samples. The Express Gene SARS-CoV2 RT-PCR Diagnostic Panel is authorized for COVID-19 detection by the FDA (EUA200423). To prevent contamination, all reagents were prepared in a PCR amplicon-free workstation. All RNA samples and components were kept on ice during use. The TaqPad Combo kit was used for detection of N gene, S gene and ORF1ab targets on COVID-19 tests. One Positive Control (1 × 104 copies/μL) and one Negative Control were included on the RT-PCR plate. RNA samples were thawed on ice, then 2.5 microliter RNA samples containing 15 ng total RNA were used for RT-PCR master mix together with primer and probes. One-step RT-PCR reaction mix (ThermoFisher) was used for RT-PCR reaction. All samples were run in triplicate. The Express Gene SARS-CoV2 RT-PCR Diagnostic Panel can detect quantities as low as 2 copies of N gene, 4 copies of ORF1ab, and 10 copies of S gene from COVID-19 viral genome per reaction volume. This information is part of our approved FDA Emergency Use Authorization (EUA200423) and can be found on the FDA website.

### Statistical Analysis

Categorical variables were presented as numbers and percentages and examined with the Mantel-Haenszel test. Continuous variables were compared using Student T-test (normally distributed) and Wilcoxon rank-sum (non-normally distributed) variables. A univariate analysis was subsequently performed to compare differences in categorical and continuous variables for each outcome. To determine risk factors for each outcome, a multivariable analysis was performed using a stepwise logistic regression model. Cox regression was performed to determine risk factors associated with in-hospital mortality. Clinically relevant factors were found based on our and other studies and added into the models ^18-21^. We used the Charlson comorbidity index because we expected a low number of outcomes for each outcome variable and were able to include only a limited number of potential confounders from the comorbidities in the model (CCI) ^22^. The statistical significance was accepted when the p value was less than 0.05. Statistical analysis was performed using IBM SPSS version 26.

## Results

A total of 400 patients were diagnosed with COVID19 infection during the study period. The demographic and clinical data of patients are reported in supplementary Table S1. Emergency physicians admitted 248 (62%) of the patients and the rest were discharged.

Among the patients who were admitted to the hospital, the mean (SD) age was 63 (17.2) years old and 141 (57%) were male, 147 (61%) were White, while 65 (44.4%) of them were White Hispanic. Sixty-two (24.5%) of admitted patients were African American and the rest multiracial.

During hospitalization, 76 (30%) patients were admitted to the MICU. In univariant analysis, patients were found to be more likely to be admitted to the MICU if they were on chemotherapy, had consolidation or bilateral opacities in chest images, or had laboratory findings of higher leukocyte counts, lower platelet counts, higher levels of BUN, serum IL6, CRP, LDH, or lower serum albumin levels(Table S2). Patients presenting complaints of dyspnea, fever, abdominal pain (but not chest pain) were more likely to be admitted to the MICU (Tables S3). The mean CCI was higher in patients admitted to the MICU compared to non-MICU patients, 3.67 vs 2.99 (P=0.047), respectively. Patients with diabetes mellitus (OR=2.4) had higher risk for admission to MICU. Patients admitted to the MICU had higher odds of in-hospital death (OR = 7.85). Among CRP levels, IL6 levels, CCI, and dyspnea, only dyspnea was found to be independently associated with increased risk of admission to the MICU, using the multivariable logistic regression model. (Table 1).

**Table 1.**
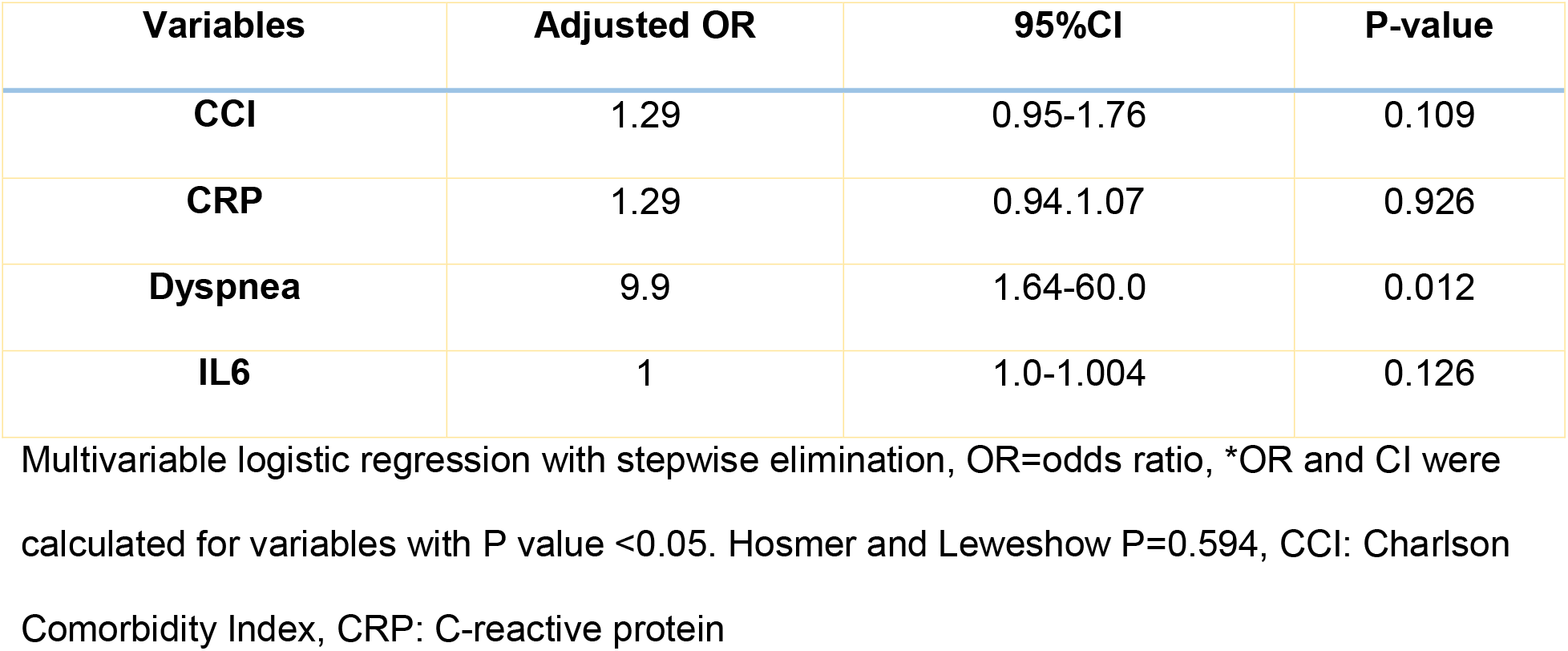
Multivariable regression model for MICU admission for patients with COVID19

A total of 30 (26%) patients died during hospitalization. Univariant analysis associated the following with higher mortality: advanced age; nursing home residency; higher CCI; a history of HTN, HF, CAD, AMS, or DM; use of ACE2 medications before admission; presentation with chest images containing consolidations with high levels of WBC, IL6, BUN, LDH; admission to the MICU; and need for noninvasive ventilation (NIV) (Tables S4, S5). The Cox regression model for in-hospital mortality showed that between CCI, high concentrations of LDH and IL6, using ACE inhibitors before admission, only CCI and IL6 were independently associated with in-hospital mortality (Table 2).

**Table 2.**
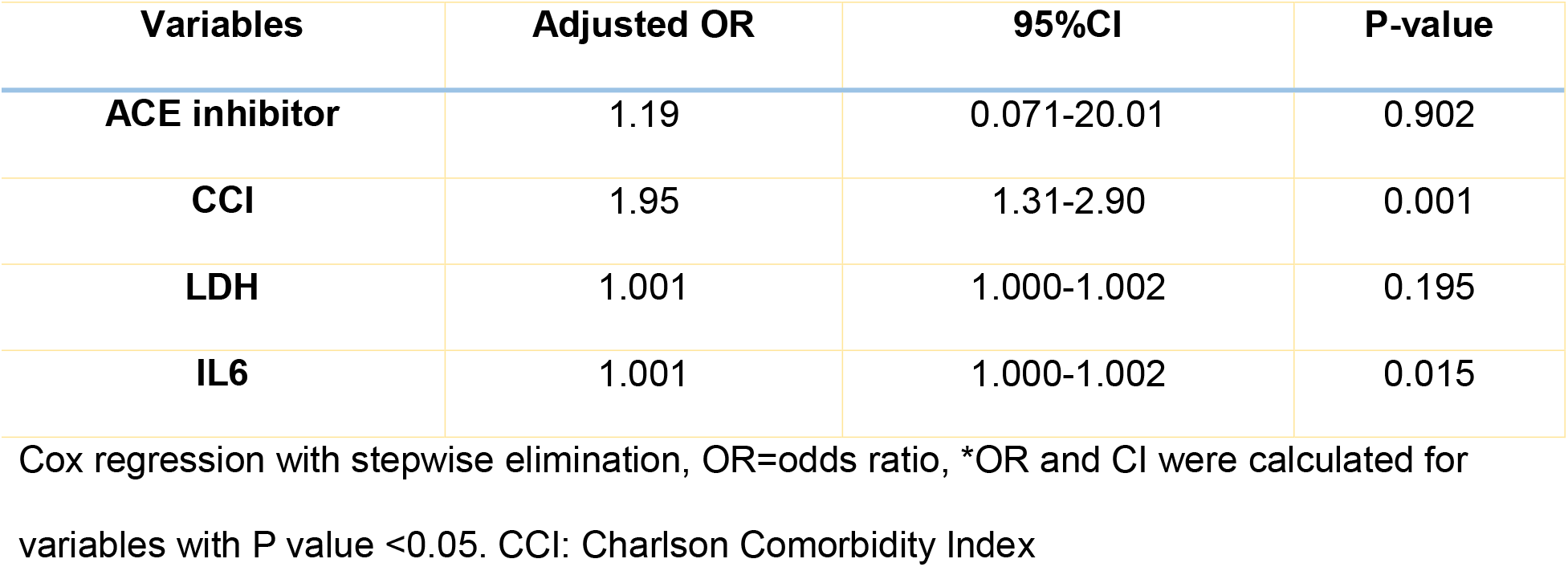
Cox regression model for in-hospital mortality for patients with COVID19

| **Variables** | **Adjusted OR** | **95%CI** | **P-value** |
| --- | --- | --- | --- |
| **ACE inhibitor** | 1.19 | 0.071-20.01 | 0.902 |
| **CCI** | 1.95 | 1.31-2.90 | 0.001 |
| **LDH** | 1.001 | 1.000-1.002 | 0.195 |
| **IL6** | 1.001 | 1.000-1.002 | 0.015 |

Cox regression with stepwise elimination, OR=odds ratio, \*OR and CI were calculated for variables with P value <0.05. CCI: Charlson Comorbidity Index

### Serum IgG analysis

Out of the 248 patients admitted with COVID-19, 100 consented to participate in the serology study. The mean (SD) age was 63 (16.9) with a median of 64, and 49 (49.5%) were male. 61 (62%) were White, while 50 (82%) were White Hispanic.

Anti-N protein IgG was positive in 55 (55%) patients at the time of admission. Tables 3 and 4 show the demographic and clinical data of the study population.

**Table 3.** Demographic characteristics of patient with high S/Co ratio for IgG against N protein

| Variables | S/Co ratio>1.52 |  | P value, OR*, CI* |
| --- | --- | --- | --- |
|  | Yes<br>Number (%) | No<br>Number (%) |  |
| Male | 15 (30.0) | 10 (20.4) | 0.274 |
| Age (years) M±SD | 63.24 ± 16.95 | 62.64 ± 15.67 | 0.876 |
| Height (cm) M±SD | 164.24 ± 21.83 | 166.48 ± 8.71 | 0.620 |
| Weight (lb.) M±SD | 169.94 ± 35.52 | 176.60 ± 45.18 | 0.457 |
| Patient delay (days) | 61 (3.85 ± 3.40) | 22 (5.68 ± 4.28) | 0.047 |
| Doctor delay (days) | 60 (3.57 ± 2.90) | 21 (5.24 ± 4.47) | 0.122 |
| Current smoker | 1 (16.7) | 24 (25.8) | 0.621 |
| Current alcohol ingestion | 7 (26.9) | 18 (24.7) | 0.819 |
| Current Marijuana ingestion/inhalational | 1 (50.0) | 24 (24.7) | 0.438 |
| Nurse Home | 4 (30.8) | 20 (24.4) | 0.624 |
| **Medication intake prior hospital admission** |  |  |  |
| Chemotherapy | 1 (20.0) | 22 (25.3) | 0.791 |
| Inhaled steroid | 1 (8.3) | 22 (27.8) | 0.178 |
| Ace inhibitors | 1 (6.3) | 22 (28.9) | 0.089 |
| Angiotensin Receptor Blockers | 2 (14.3) | 21 (26.6) | 0.335 |
| Statins | 6 (18.2) | 17 (27.9) | 0.300 |
| **CT or Chest Xray findings obtained after hospital Admission** |  |  |  |
| Ground glass opacities | 5 (21.7) | 20 (27.0) | 0.613 |
| Consolidation | 2 (13.3) | 23 (28.0) | 0.244 |
| Bilateral infiltrates | 18 (34.6) | 7 (15.6) | 0.036, 2.87 (1.07-7.72) |
| **Laboratory Findings within 48 hours of hospital admission** |  |  |  |
| Leukocytes ($10^3/\mu\text{L}$) | 74 ($7.93 \pm 5.27$) | 25 ($9.32 \pm 5.02$) | 0.254 |
| Neutrophils ($10^3/\mu\text{L}$) | 74 ($8.89 \pm 13.52$) | 25 ($7.89 \pm 4.58$) | 0.718 |
| Lymphocytes ($10^3/\mu\text{L}$) | 74 ($2.42 \pm 4.90$) | 25 ($1.06 \pm 0.67$) | 0.022 |
| Eosinophils ($10^3/\mu\text{L}$) | 72 ($0.07 \pm 0.18$) | 23 ($0.03 \pm 0.08$) | 0.318 |
| Hematocrit (%) | 74 ($35.40 \pm 7.63$) | 24 ($38.98 \pm 6.81$) | 0.035 |
| Platelet count ($10^3/\mu\text{L}$) | 74 ($219.39 \pm 100.75$) | 24 ($248.25 \pm 137.06$) | 0.349 |
| WBC>10000 | 8 (34.8) | 17 (22.4) | 0.234 |
| WBC<4500 | 4 (17.4) | 21 (27.6) | 0.327 |
| Interleukin-6 (pg/mL) | 24 ($288.25 \pm 707.60$) | 13 ($674.76 \pm 1097.43$) | 0.201 |
| IL6>186 | 5 (55.6) | 8 (28.6) | 0.150 |
| C-Reactive Protein (mg/dL) | 66 ($10.53 \pm 14.12$) | 24 ($14.88 \pm 11.39$) | 0.178 |
| CRP>15 | 9 (37.5) | 15 (22.7) | 0.165 |
| Erythrocyte sedimentation rate (mm/hr) | 12 ($50.67 \pm 25.18$) | 4 ($59.75 \pm 40.97$) | 0.600 |
| Blood urea nitrogen (mg/dL) | 74 ($22.90 \pm 21.19$) | 25 ($23.64 \pm 27.38$) | 0.889 |
| Creatinine (mg/dL) | 74 ($1.73 \pm 2.67$) | 25 ($1.20 \pm 0.96$) | 0.333 |
| Ferritin (ng/mL) | 64 ($1171.09 \pm 2343.46$) | 23 ($1117.13 \pm 895.65$) | 0.915 |
| Procalcitonin (ng/mL) | 58 ($1.33 \pm 3.51$) | 21 ($1.68 \pm 6.12$) | 0.752 |
| Lactate (mmol/L) | 61 ($1.99 \pm 1.71$) | 21 ($2.51 \pm 2.83$) | 0.322 |
| Troponin (ng/mL) | 43 ($0.03 \pm 0.06$) | 18 ($105.72 \pm 448.29$) | 0.331 |
| Creatine kinase (U/L) | 38 ($199.71 \pm 357.05$) | 17 ($425.75 \pm 504.77$) | 0.108 |
| Lactate dehydrogenase (U/L) | 61 ($398.10 \pm 324.86$) | 24 ($459.38 \pm 195.58$) | 0.391 |
| Fibrinogen (mg/dL) | 7 (520.43 ± 186.21) | 1 (474.00 ± N/A) | 0.823 |
| ALT (U/L) | 69 (70.04 ± 204.74) | 23 (69.35 ± 64.73) | 0.987 |
| AST (U/L) | 69 (128.81 ± 642.59) | 23 (85.74 ± 86.31) | 0.750 |
| Albumin (mg/dL) | 69 (3.67 ± 0.50) | 23 (3.35 ± 0.53) | 0.017 |
| Bilirubin (mg/dL) | 68 (0.65 ± 1.42) | 23 (0.63 ± 0.48) | 0.949 |
| D-dimer (ug/mL) | 47 (7.81 ± 39.07) | 18 (5.62 ± 7.59) | 0.815 |

\*OR and CI were calculated for categorical variables with P value <0.05, M±SD: Mean±
standard deviation, ALT: Alanine aminotransferease, AST: Aspartate aminotransaminase

**Table 4.** Comorbidities and clinical outcomes of patient with high S/Co ratio for IgG against N protein

| **Variables** | **S/Co ratio>1.52** |  | **P value, OR\*, CI\*** |
| --- | --- | --- | --- |
| **Comorbidities** | **Yes**<br>**Number (%)** | **No**<br>**Number (%)** |  |
| **Asthma** | 1 (8.3) | 23 (28.8) | 0.165 |
| **Pulmonary embolism** | 1 (25.0) | 24 (26.7) | 0.941 |
| **COPD** | 3 (30.0) | 22 (25.9) | 0.780 |
| **Emphysema** | 1 (50.0) | 24 (26.1) | 0.468 |
| **Chronic heart failure** | 1 (20.0) | 24 (27.0) | 0.733 |
| **Coronary Artery Disease** | 1 (12.5) | 24 (27.9) | 0.363 |
| **Atrial fibrillation** | 3 (33.3) | 22 (25.6) | 0.617 |
| **Hypertension** | 14 (24.6) | 11 (28.2) | 0.690 |
| **Stroke** | 1 (20.0) | 23 (25.8) | 0.772 |
| **Dementia** | 3 (30.0) | 22 (25.6) | 0.764 |
| **Chronic Renal Failure** | 1 (10.0) | 23 (27.4) | 0.259 |
| **Liver Disease** | 1 (33.3) | 24 (26.4) | 0.789 |
| **Diabetes Mellitus** | 7 (22.6) | 18 (28.1) | 0.74 |
| **Clinical Symptoms** |  |  |  |
| **Cough** | 20 (33.3) | 5 (13.9) | 0.041, 3.10 (1.04-9.18) |
| **Sputum** | 4 (57.1) | 21 (24.4) | 0.078, 4.12 (0.85-19.95) |
| **Chest pain** | 4 (40.0) | 21 (25.3) | 0.329 |
| **Dyspnea** | 21 (35.0) | 4 (10.5) | 0.010, 4.57 (1.42-14.65) |
| **Hemoptysis** | 2 (66.7) | 23 (24.7) | 0.148 |
| **Fever** | 19 (29.7) | 6 (18.2) | 0.224 |
| **Chills** | 9 (31.0) | 16 (24.6) | 0.516 |
| **Headache** | 4 (36.4) | 21 (25.3) | 0.439 |
| **Myalgia** | 3 (15.8) | 22 (29.7) | 0.230 |
| **Abdominal pain** | 2 (15.4) | 23 (28.8) | 0.323 |
| **Diarrhea** | 6 (30.0) | 19 (25.7) | 0.698 |
| **Nausea or vomiting** | 6 (40.0) | 19 (24.4) | 0.217 |
| **Altered Mental Status** | 3 (21.4) | 22 (26.5) | 0.689 |
| **Outcomes** |  |  |  |
| **Hospital LOS>16 days** | 11 (47.8) | 14 (18.4) | 0.006, 4.060 (1.48-11.06) |
| **MICU stay>8 days** | 10 (71.4) | 1 (7.1) | 0.004, 32.50 (3.12-337.81) |
| **MICU Admission** | 11 (39.3) | 14 (19.7) | 0.047, 2.63 (1.01-6.86) |
| **MICU Death** | 3 (60.0) | 22 (23.4) | 0.092, 4.90 (0.77-31.27) |
| **Hospital death** | 3 (30.0) | 22 (24.7) | 0.716 |
| **MICU stay (days)** | 17 (9.18 ± 10.35) | 11 (16.82 ± 10.12) | 0.065 |
| **Hospital stay (days)** | 74 (11.39 ± 10.33) | 25 (17.68 ± 13.53) | 0.041 |

\*OR and CI were calculated for variables with P value <0.05. S/Co: Signal to cut off, COPD:
Chronic obstructive pulmonary disease, LOS: Length of stay,

In univariate analysis, an S/Co ratio greater than 1.5 was found to be associated with increased likelihood of admission with symptoms of dyspnea (p=0.01, OR= 4.57) and cough (p=0.041, OR= 3.10). Patients with S/Co ratio >1.5 were more likely to have bilateral infiltrates on admission (p=0.036, OR= 2.87). There was a significant difference (p=0.035) in mean hematocrit percentage in patients with an S/Co ratio > 1.5 (M= 35.40 ± 7.63) vs those with lower S/Co ratio (M=38.98 ± 6.81). There were also significant (p=0.017) differences in albumin levels for patients who had an S/Co ratio > 1.5 (3.67 ± 0.50 g/dl) and those who did not (3.35 ± 0.53 g/dl). There were also significant (p=0.022) differences in mean lymphocyte levels for patients with an S/Co > 1.5 (2.42 ± 4.90 ×10^3^ cells/ml) and patients who did not have an S/Co > 1.5 (1.06 ± 0.67 × 10^3^ cells/ml). S/Co ratio > 1.5 was associated with increased likelihood of ICU admission (p=0.047, OR=2.63 days). S/Co >1.5 was also associated with increased likelihood of staying in the hospital for more than 16 days (p=0.006, OR= 4.06) and MICU stays of more than 8 days (p=0.004, OR= 32.50) (Table 4). There was a significant correlation between IgG S/Co ratio and days in MICU as shown in Figure 2.

**Figure 2.**
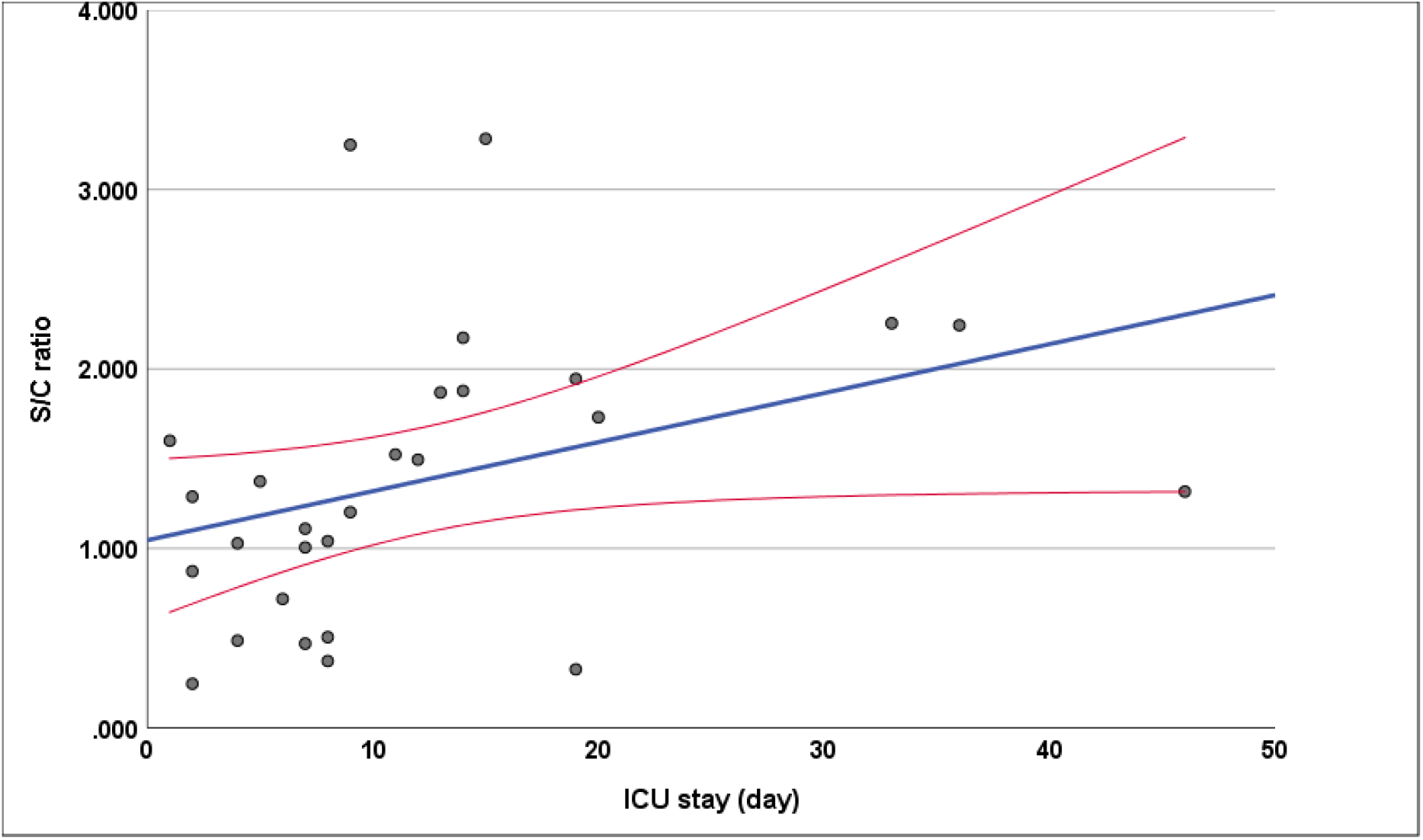
Shows a correlation between SARS-CoV-2 IgG for N protein and MICU admission duration. R=0.554 and P-Value = 0.002

The multivariable logistic regression model for MICU admission showed that between African American, CCI, lymphocyte counts, and S/Co ratio> 1.5, only S/Co ratio were independently associated with MICU admission (Table 5). In multivariable analysis for hospital stays of more than 16 days, S/Co ratio>1.5 (p=0.047, OR:2.63) was again the only independent factor among factors the mentioned variables (p=0.006 OR:4.1) (Table 6).

**Table 5.**
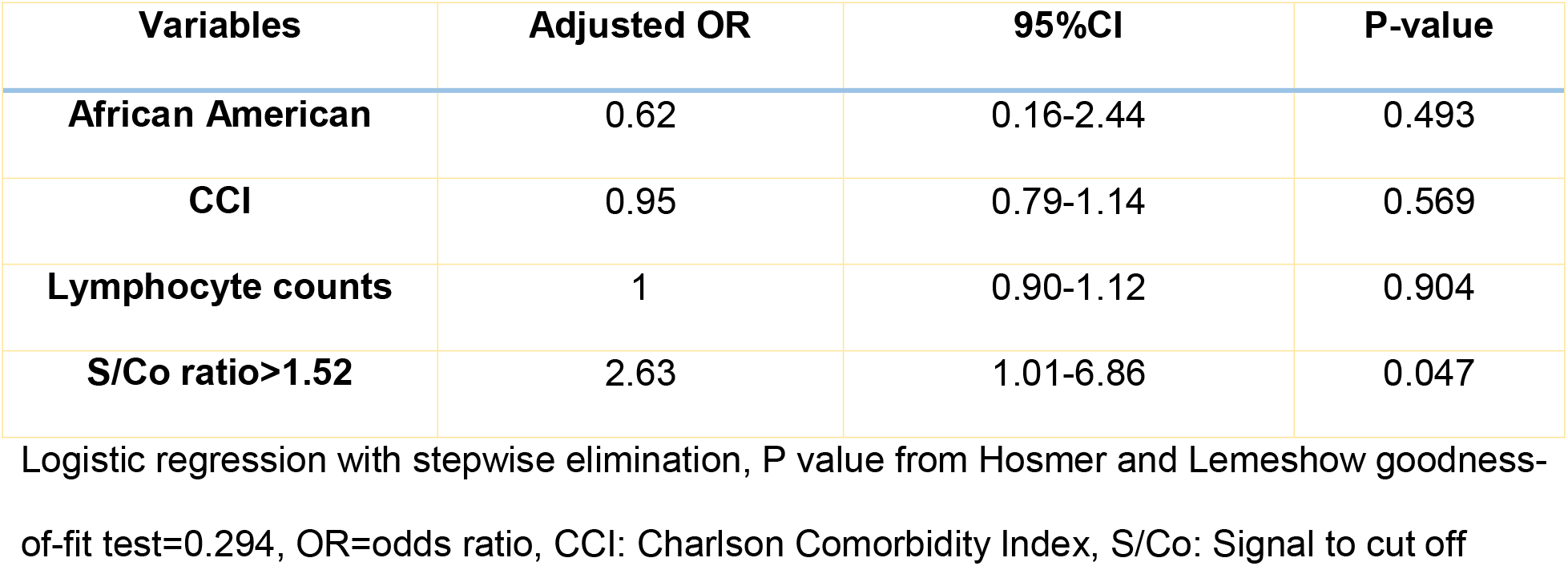
Multivariable logistic regression model for MICU admission days for patients with high IgG against N-protein

**Table 6.** Multivariable logistic regression model for Hospital stay>16 days for patients with high IgG against N-protein

| **Variables** | **Adjusted OR** | **95%CI** | **P-value** |
| --- | --- | --- | --- |
| **African American** | 0.88 | 0.21-3.61 | 0.855 |
| **CCI** | 1 | 0.82-1.23 | 0.985 |
| **Lymphocyte counts** | 0.931 | 0.74-1.16 | 0.528 |
| **S/Co ratio>1.5** | 4.1 | 1.49-11.1 | 0.006 |

Logistic regression with stepwise elimination, P value from Hosmer and Lemeshow goodness-of-fit test=0.887, OR=odds ratio. CCI: Charlson Comorbidity Index, S/Co: Signal to cut off

### Whole blood RT-PCR

The RT-PCR reaction indicated undetermined levels of N gene, S gene, and ORF1ab nucleotides sequences on prepared RNA samples from subjects with negative, low, and high concentrations of IgG. RT-PCR data is shown in the supplemental Excel file.

## Discussion

This study indicates that elderly males with COVID19 and a higher number of comorbidity scores were more likely to be admitted to the hospital. Among hospitalized patients, we found only high levels of dyspnea independently predicted MICU admission. We also demonstrated that higher CCI and higher IL6 levels were independently associated with in-hospital mortality. COVID19-confirmed patients admitted to the hospital for hypoxemia are likely to have developed a high concentration of IgG against the N protein of SARS-CoV2. High IgG levels are associated with a higher risk of admission to the MICU and a more extended stay in the hospital. We also found no evidence of viremia in our hospitalized COVID19 patients.

The present investigation is the first to report an association of high concentration of IgG against the N protein with poor outcome in COVID19. Prior studies reported detectable levels of total IgG in COVID19 patients in the first week of the disease^23,24^. We found that a high concentration of IgG against N-protein caused a 3-fold increase in risk of admission to the MICU. It could be theorized that N protein IgG may favor a higher inflammatory response during infection.

Neutralizing Abs are usually produced against viral entry proteins ^25-27^. The IgG against intact S protein or the S1 subunit of S protein is accepted as a neutralizing antibody against SARS and COVID19 ^28-30^. The IgG targeting S protein is likely protective, and it has become the primary target for COVID19 vaccine development ^31^. There is no evidence to suggest that S protein IgG (via vaccine or passive IgG infusion) induces lung pathology via the ADE effect ^32^. Although the potential role of ADE in COVID19 remains unknown, the current study suggests that the IgG levels of non-neutralizing nucleoprotein may be a reason for poor outcomes in COVID19 subjects. The high levels of non-neutralizing IgG for N protein might play a role as a Trojan Horse for SARS-CoV2 and facilitate virus entry to immune cells with attachment to fragment crystallizable (Fc) regions of lymphocytes, dendritic cells, macrophages, natural killer cells, and even platelets. The infected immune cells respond with cytokine production, and autophagy ^33^. Interestingly, a higher rate of reported pulmonary embolism and thrombosis in patients with severe COVID19 ^34-36^ may be attributed to platelet activation after direct infection with the virus and secondary autophagy ^37,38^.

Another possible pathologic mechanism could be antibody-mediated cellular cytotoxicity in infected cells expressing N protein particles in the cell wall. Natural killer cells, neutrophils, and macrophages interact with IgG and eliminate the target cells ^39-41^. Immune complex development against N protein is a fascinating potential mechanism. Berger and his colleagues showed that the immune complex induces IL6 secretion by immune cells and activates a network of proinflammatory cytokines and profound systemic inflammatory response ^42^. Immune complex against N protein may contribute to the hyperinflammatory response that has been reported in COVID19 ^43^.

The role of ADE in the pathogenesis of dengue has been reviewed previously ^44^. In animal models studying the pathogenesis of the dengue virus, ADE was associated with higher viremia ^45-47^. However, the current study suggests that the pathogenesis of SARS-CoV2 is not correlated with higher ADE-associated viremia. Our observation may suggest that SARS-CoV2 might stay in the lymphatic system and avoid the bloodstream. Further investigation is urgently needed to confirm our findings.

The major limitation of our study is that we did not measure total non-neutralizing IgG and non-neutralizing anti-S and other proteins. In addition, we were unable to show the effect of IgG targeting N protein on mortality of COVID19 patients in the multivariable analysis due to sample size limitations. This study has not yet been validated in another cohort of COVID19 patients.

Further investigation is urgently needed to assess the pathologic mechanisms of IgG for N-protein in severe COVID19. In particular, a larger multicentric study should be conducted to investigate the role of IgG targeting N protein on COVID19 mortality. A better understanding of the role of ADE in platelet activation may shed light on the hypercoagulable state in COVID19 and generate new therapeutic modalities.

In summary, this study recommends that during the initial assessment of patients with COVID19, IgG targeting N-protein of SARS-CoV2 should be included among the measures. The high concentration of this immunoglobulin may predict poor outcomes, although further validation is needed.

## Data Availability

Data available on request from the authors.

## Acknowledgments

Authors would like to acknowledge the nursing staff of the COVID19 units of University of Miami for such a wonderful collaboration with our study team. Authors would like to thank Gary and Barbara Paige for their donations to our research group.

## Author contributions

MM conceptualized the study. MM and CZ designed the experiments. CZ, RT, and EC conducted IgG experiments. MM and CZ analyzed the data. MB, CSS, JNM, JRDLF, MM, DG, SP, MVPB, SH, MM, MSG, SK, KHMS, HA performed clinical data collections. FM and MF performed RT-PCR tests from blood samples. MM wrote the manuscript. All authors reviewed and edited the manuscript.

